# A sex-stratified analysis of the genetic architecture of human brain anatomy

**DOI:** 10.1101/2023.08.09.23293881

**Authors:** Rebecca Shafee, Dustin Moraczewski, Siyuan Liu, Travis Mallard, Adam Thomas, Armin Raznahan

## Abstract

Large biobanks have dramatically advanced our understanding of genetic influences on human brain anatomy. However, most studies have combined rather than compared males and females - despite theoretical grounds for potential sex differences. By systematically screening for sex differences in the common genetic architecture of > 1000 neuroanatomical phenotypes in the UK Biobank, we establish a general concordance between males and females in heritability estimates, genetic correlations and variant-level effects. Notable exceptions include: higher mean h^2^ in females for regional volume and surface area phenotypes; between-sex genetic correlations that are significantly below 1 in the insula and parietal cortex; and, a male-specific effect common variant mapping to *RBFOX1 -* a gene linked to multiple male-biased neuropsychiatric disorders. This work suggests that common variant influences on human brain anatomy are largely consistent between males and females, with a few exceptions that will guide future research as biobanks continue to grow in size.

## Introduction

Our understanding of genetic influences on human brain anatomy has expanded rapidly in recent years (Grasby et al., 2020; Satizabal et al., 2019; Smith et al., 2021) due to the availability of combined neuroimaging and genetic information in large datasets such as the UK Biobank [UKB, (Sudlow et al., 2015)] and international consortia [e.g. ENIGMA (Thompson, 2020)]. Collectively, the rapidly growing number of studies in such datasets has: established the high heritability of many neuroanatomical phenotypes; revealed regional variation in the heritability and genetic architecture across different features of the brain; identified sets of genetic variants that shape different global and regional aspects of brain anatomy; and, established overlaps between the genetic determinants of brain anatomy and risk for brain-based neuropsychiatric disorders (Elliott et al., 2018; Grasby et al., 2020; Smith et al., 2021). However, with few exceptions (Smith et al. 2021; Zhao et al. 2019), this growing and impactful literature has typically combined males and females rather than directly compared genetic influences on brain anatomy between males and females.

Several observations strongly motivate comparing the genetic architecture of neuroanatomical variation in males and females. Structural magnetic resonance imaging (sMRI) studies of brain anatomy have identified several reproducible sex differences in brain anatomy including greater mean total brain volume in males than females (Ritchie et al., 2018; Williams et al., 2021a) which survives statistical correction for sex-differences in height (Williams et al., 2021b), and sex-differences in regional brain anatomy above and beyond these differences in overall brain size (DeCasien et al., 2022; Liu et al., 2020; Lotze et al., 2019; Williams et al., 2021a). If these phenotypic sex differences partly reflect sex-specific biological influences on brain development, then this would provide an opportunity for sex-biased genetic influences on brain anatomy. In support of this reasoning, a large corpus of experimental research in animal models indicates that several canonical sex-differences in regional anatomy of the mammalian and rodent brain are indeed determined by sex-specific influences of gonadal steroids and sex chromosomes (Corre et al., 2016; DeCasien et al., 2022; McCarthy, 2020; McCarthy et al., 2012, 2017; Neufang et al., 2008; Premachandran et al., 2020; Vousden et al., 2018). For example, because several regions of sex-biased brain volume in rodents are established by male-specific influences of testosterone (via aromatization to estradiol) on apoptosis (Wright et al., 2010) - genetic variation in the strength of these influences would be predicted to modulate interindividual variation of region size in males more prominently than females. In a similar fashion, all placental mammals show sex differences in the dosage of X and Y chromosomes (males XY and females XX) - which contain genes that are known to influence regional brain anatomy (Guma et al., 2022; Mallard et al., 2021; Warling et al., 2021) and thereby introduce sex-specific genetic sources of neuroanatomical variation. Finally, the potential for sex differences in genetic influences on neuroanatomical variation is also suggested by the observation that neuroanatomical correlates of several heritable neuropsychiatric disorders have been reported to differ between males and females (Guma et al., 2017; Supekar et al., 2022), which could arise if disease-relevant genetic variants differentially influenced brain anatomy as a function of sex. Despite these numerous grounds for hypothesizing sex differences in the architecture of genetic influences on brain anatomy - we have so far lacked any direct tests for such differences in humans.

Here, we use the UK Biobank sample (UKB, males = 14534, females = 16294, **Supplementary Table S1)** to systematically compare the genetic architecture of neuroanatomical variation in males and females (**Figure 1**). We examine 1106 phenotypes including: 1080 regional measures of cortical anatomy encompassing estimates of gray matter volume (GMV), surface area (SA) and cortical thickness (CT) for 360 regions of interest [(Glasser et al., 2016) each corrected for the corresponding global brain phenotype]; 23 subcortical structure volumes (**Supplementary Table S1,** corrected for total brain volume), and 3 global measures (mean cortical thickness, total cortical surface area and total gray matter volume). We distinguish between these different morphometric properties of the brain because they are known to show distinct genetic architectures (Elliott et al., 2018; Grasby et al., 2020; Smith et al., 2021) and varying mean differences between males and females (Liu et al., 2020; Williams et al., 2021a). We first compare the total SNP-heritability (h^2^ autosomal and X-chromosomal) of each phenotype between males and females. Next, we evaluate genetic correlations (r_g_) between males and females for each phenotype screening for any potential instances where this correlation differs from 1. Finally, we carry out sex-stratified genome wide association analyses (GWAS) for each phenotype to test for any genetic variants with significantly different effects in the two sexes.

**Figure 1:**
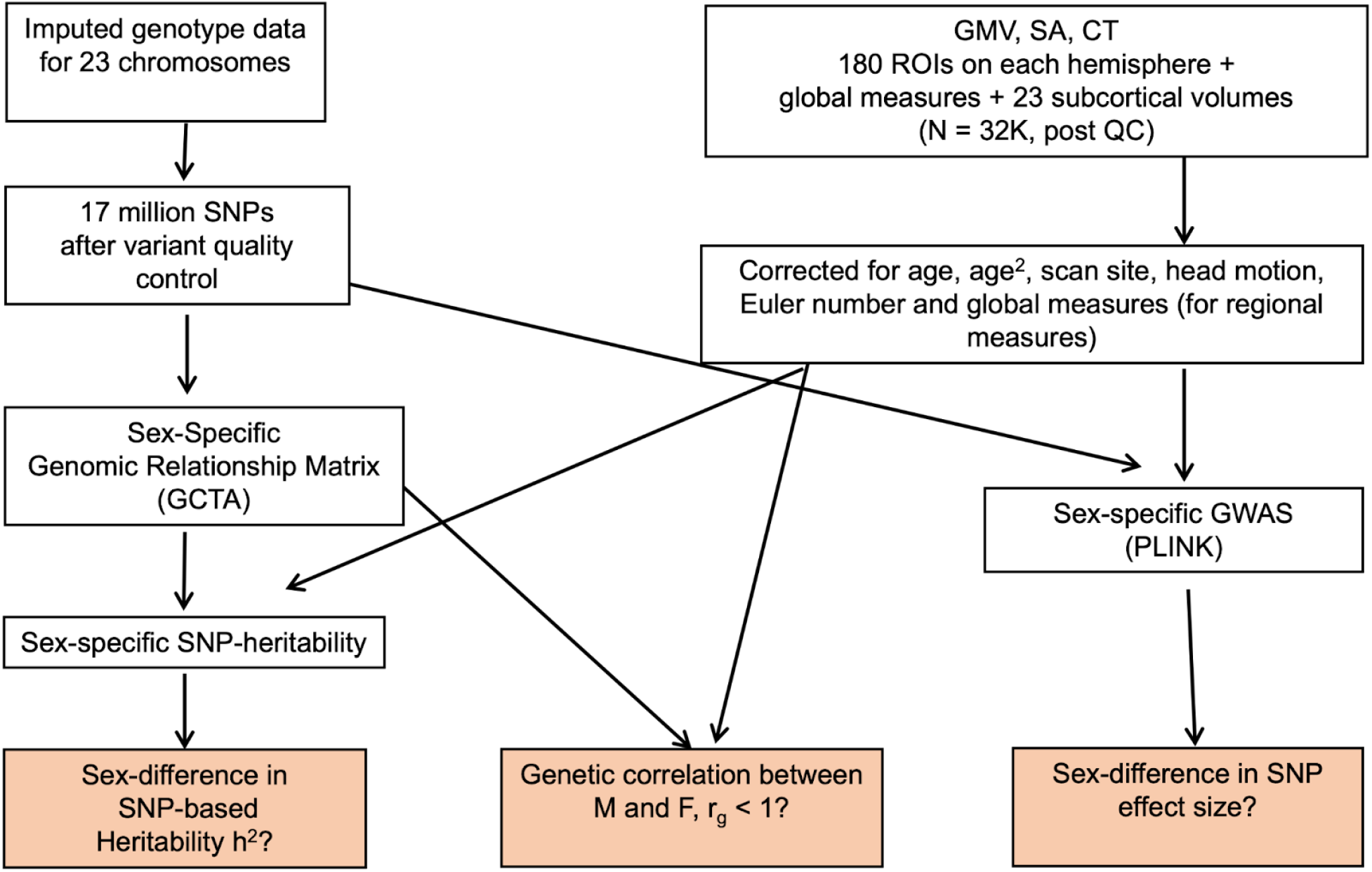
Outline of analysis of sex-difference in the genetic architecture of brain MRI phenotypes in the UK Biobank sample.

Our systematic screen finds that the genetic architecture of neuroanatomical variation in humans is broadly congruent between males and females, but also highlights three notable sex differences. First, there is a general tendency across all brain regions for females to show higher mean SNP-based heritability (mean h^2^) for cortical GMV and SA than males. Second, we find that the strength of genetic correlations (r_g_) between males and females varies substantially across the cortical sheet and falls significantly below 1 for isolated regions of the parietal cortex and insula). Finally, after stringent control for multiple comparisons across all brain regions we find statistically significant sex-differences in the phenotypic effects of a common variant mapping to *RBFOX1* - a known risk gene for sex-biased neuropsychiatric and neurodevelopmental disorders like ASD and schizophrenia (Cross-Disorder Group of the Psychiatric Genomics Consortium, 2019; Fogel et al., 2012). Across all phenotypes, 517 more genes show evidence for sex-biased SNP effects at a genome-wide level of statistical significance (p < 5e-8), several of which are likely to reach multiple testing corrected statistical significance with expanding sample sizes (Visscher et al., 2017). Taken together, these results provide a benchmark view of sex-differences in the genetic architecture of human brain anatomy which points toward general convergence between males and females, but also highlights important instances of divergence which warrant repeated investigation as biobank sample sizes increase.

## Results

### Sex-difference in SNP-based heritability

Most brain phenotypes are significantly heritable and it is possible to estimate the fraction of the phenotypic variance that is captured by genotyped SNPs using the genomic relationship matrix (GRM) approach. Using GCTA (Yang et al., 2011) we constructed sex-specific autosomal and X-chromosomal GRMs from which SNP-heritability (h^2^) was estimated in each sex for: 180 regional measures (HCP parcellation, Glasser et al., 2016) of GMV, SA and CT in each hemisphere; 23 subcortical volumes; and, 3 global measures (total 1106 phenotypes).

Figure 2 shows the spatial maps of total h^2^ for cortical GMV, SA and CT in each sex (Figure 2A, **B** and **C**, respectively). Separate autosomal and X-chromosomal contributions to h^2^ can be found in the **Supplementary Tables** (**Tables S2, S3, S4**). X-linked heritability was estimated using the best-fit dosage compensation models (full dosage compensation/no dosage compensation/equal variance) from Mallard et al., (2021) for each phenotype. The spatial variation in h^2^ estimates was qualitatively similar between males and females for regional GMV, SA and CT and the between-sex correlation in h^2^ across all brain regions was high for all three phenotypic classes (r = 0.82 for GMV, r = 0.83 for SA and r = 0.67 for CT). Comparison of male and female h^2^ for each phenotype (using the estimated h^2^ and its standard error for each sex, **Methods**) resulted in no significant regional difference after correction for multiple testing across all cortical regions (all p > 1.4e-4, **Supplementary Tables S2, S3, S4).** We observed some regional variation in the magnitude of sex-differences in h^2^ however (Figure 2G**, H and I**), which was unrelated to the magnitude of phenotypic sex differences (calculated as described in **Methods**) for GMV and SA, but showed a statistically significant and weakly negative correlation with regional variation in phenotypic sex-differences for CT (r = -0.14, p = 0.007, **Figure S2F**). However, when considering the distribution of h^2^ across all cortical regions collectively, mean h^2^ was statistically significantly higher in females than males for both GMV and SA (paired t-test p = 8.44 e-13 for GMV and p = 1.17 e-8 for SA; Wilcoxon rank test p = 1.64e-11 for GMV, p = 1.4e-7 for SA; Figure 2D,E,F).

**Figure 2:**
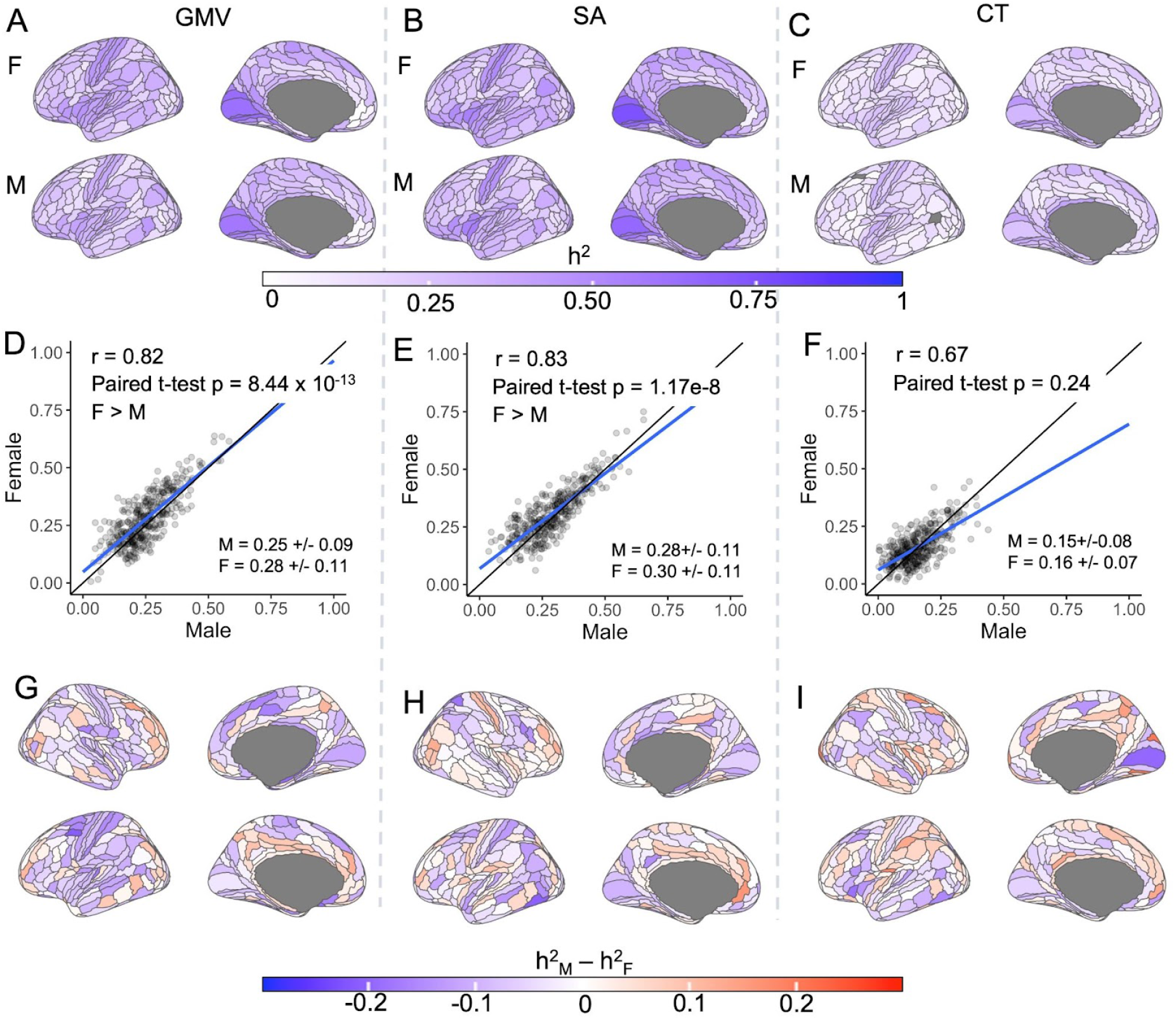
Comparing SNP-based h^2^ of regional GMV, SA and CT between males and females. (**A, B, C**): Sex-specific (M: male, F: female) spatial maps of SNP-based heritability (h^2^) of regional cortical GMV (**A**), SA (**B**) and CT (**C**). Only left hemispheres are shown - results for both hemispheres can be found in Fig S1. (**D, E, F**): Scatter plots of male and female h^2^ values of 360 cortical regions in the HCP parcellation (Methods) for GMV (**D**), SA (**E**) and CT (**F**), respectively - with inset statistics for the correlation in h^2^ across regions, and for the paired t-test of regional h^2^ between sexes. Solid blue line shows linear fit of the data. (**G**, **H**, **I)**: Spatial map of sex-differences in h^2^ for GMV, SA and CT, respectively. All phenotypes were corrected for corresponding global measures (mean thickness, total surface area and total brain volume).

We observed moderate to high total h^2^ for all 23 subcortical volumes and all three global measures, which did not differ significantly between the sexes (**Supplementary Tables S5,S6, Figure S3**).

Since by definition h^2^ is the ratio of genetic variance V_G_ and phenotypic variance V_P_, where V_P_ = V_G_ + V_E_, (V_E_ is the residual variance not attributable to additive genetic effects), we sought to refine the above finding - of greater mean h^2^ in females vs. males for regional GMV and SA measures - by examining the relationships between h^2^, V_G_, V_P_ and V_E_ in both sexes. Paired t-tests (as well as nonparametric Wilcoxon rank tests) between the sexes indicated higher mean V_G_, V_p_ and V_E_ in males compared to females in GMV (t-stat > 5.9 and p < 5e-9 for all three) and SA (t-stat > 7, p < 5.4e-12 for for all three), but not CT (all three t-test p > 0.05) (**Supplementary Table S7**). Thus the higher mean V_P_ in males for regional GMV and SA measures in the current analyses as well as reported previously (Ritchie et al., 2018) is accompanied by a higher mean V_G_, but for a given V_p_ these traits show a higher V_G_ in females than males which leads to a higher mean h^2^ estimate across traits (**Figure S4, Supplementary Table S8**).

### Between-sex genetic correlation in brain anatomy

For each phenotype we used the “bivariate” option in GCTA to calculate genetic correlation (r_g_) between males and females. Since low h^2^ can make the estimation of r_g_ unstable, we limited r_g_ calculations to phenotypes with h^2^ p-values of 0.05 or lower in both sexes (all subcortical and global phenotypes; 346 GMV regional phenotypes, 352 regional SA phenotypes, and 255 regional CT phenotypes). This analysis was also restricted to autosomes (covering 95% of the genome) because of the relatively small X-chromosome heritability contributions. Across cortical regions, The strength of between-sex r_g_ varied substantially across regions (Figure 3; GMV: 0.38 to 1; SA: 0.5 to 1; CT: 0.004 to 1), but median values were consistently high (GMV: 1; SA: 1; CT: 0.98). Likelihood tests to detect r_g_ values less than 1 were not significant for most regions (Figure 3**, Supplementary Tables S9, S10, S11**) – indicating broad between-sex similarity in the common genetic architecture of cortical anatomy. Only two regional phenotypes possessed between-sex r_g_ values that were statistically significantly lower than one (p < 1.4e-4, Figure 3): CT of left inferior posterior insula (PoI1, r_g_ = 0.46 +/- 0.11) and left superior parietal lobule medial Brodmann Area 5 SA (BA 5m, r_g_ = 0.50 +/- 0.11). Of note, these two phenotypes also showed statistically-significant sex-bias in mean values within the UKB - SA of medial BA 5 region was female-biased and CT of PoI1 was male-baised (**Figure S2 panels B,C**). However, the general relationship between the magnitude of phenotypic sex differences (calculated as described in **Methods**, also **Figure S2**) and r_g_ across the cortex was not significant for GMV and CT (p > 0.05), but weakly positive for SA (r = 0.20, p = 0.0001, with lower r_g_ regions being more female-biased (Figure 3).

**Figure 3:**
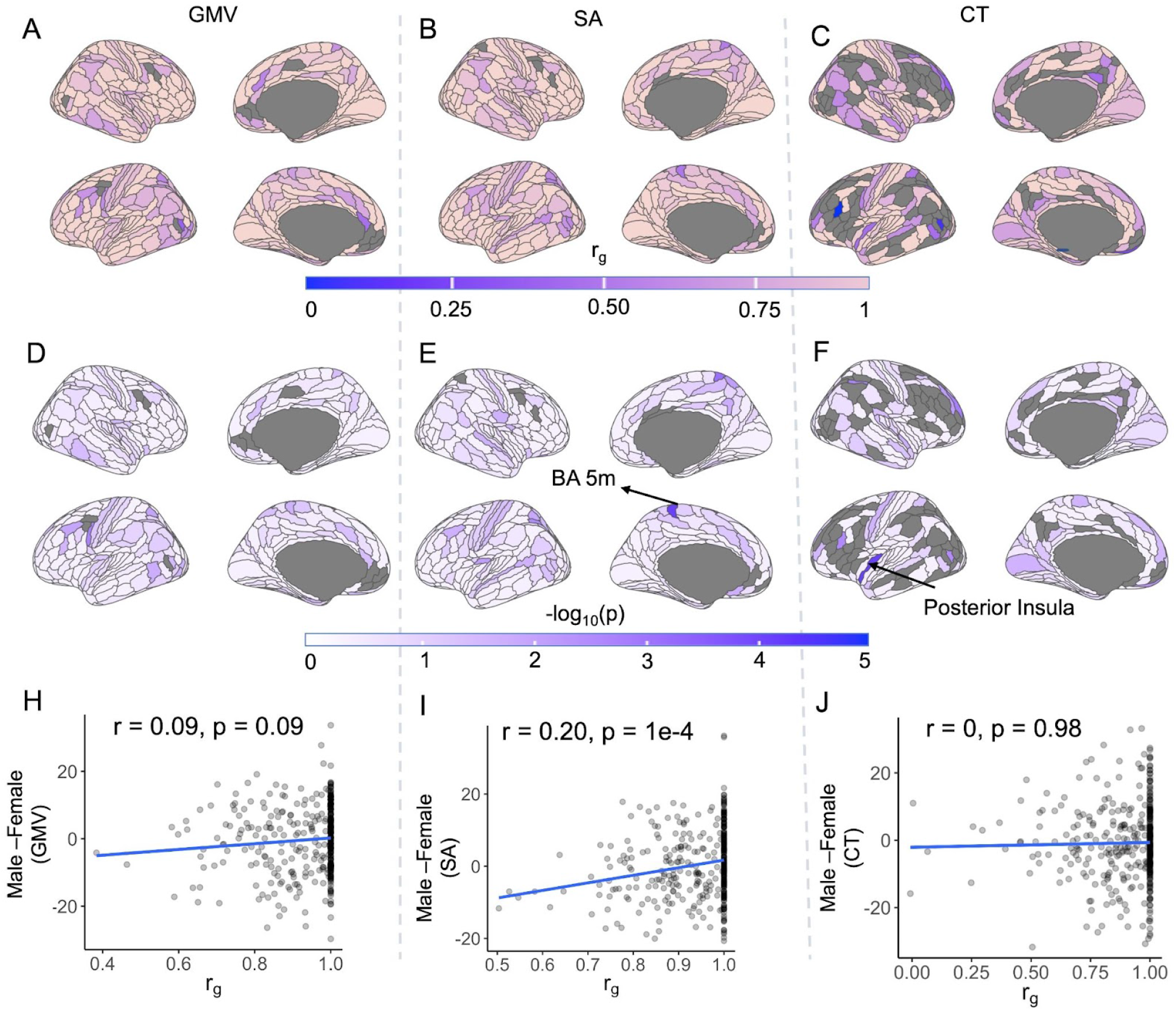
Between-sex genetic correlations (r_g_) in brain anatomy. (**A, B, C)**: Genetic correlation (r_g_) across all autosomes between males and females in regional GMV (**A**), SA (**B**) and CT (**C**) calculated using GCTA. Regions with low heritability (p > 0.05 for h^2^ in either sex) were excluded in this step and are shown in gray. (**D, E, F)**: p-values (-log10(p)) of likelihood ratio tests of r_g_ < 1 for regional GMV (**D**), SA (**E**) and CT (**F**). Only two regions showed significant r_g_ < 1 after multiple-testing correction (p < 1,4e-4): one in SA (superior parietal lobule medial Brodmann area 5, 5m, p= as 7.9 x 10^-5^, r_g_ = 0.50 +/- 0.11) and one in CT (posterior insula, PoI1, p = 7.7e-5, r_g_ = 0.46 +/- 0.11). All regional phenotypes were corrected for corresponding global measures (mean thickness, total surface area and total brain volume) prior to calculating r_g_. (**H, I, J)**: Comparison of phenotypic sex difference and genetic correlation, r_g_, in GMV (**H)**, SA (**I**) and CT (**J**) for each region of the HCP parcellation with non-negligible h^2^ in both sexes (p < 0.05). Phenotypic sex-difference is shown in t-statistics (Methods) with positive values indicating higher in males.

Between-sex r_g_ was statistically indistinguishable from 1 for all global and subcortical phenotypes (**Supplementary Tables S12, S13**). Increasing sample size in the future would help to estimate r_g_ more accurately and therefore likely bring many of the regions with between-sex r_g_ values substantially below 1 into statistical significance (e.g. - GMV of left lateral occipital lobe, LO1: r_g_ = 0.64 +/- 0.16, SA of left middle temporal visual region, MST: r_g_ = 0.62 +/- 0.18, and CT of right middle temporal gyrus, TE1p: r_g_ = 0.68 +/- 0.20).

### Sex-difference in SNP effects

We screened for individual SNPs with sex-biased effects on brain phenotypes using sex-stratified GWAS. For each SNP and each phenotype, the sex difference in effect sizes was calculated as a z-score through Equation 1 (**Methods**) using the sex-specific effect sizes and the corresponding standard errors from which a corresponding p-value was estimated assuming normal distribution in R (**Methods**). Significant sex-differences were considered at two different thresholds: (i) A “strict” threshold which accounted for multiple testing across brain regions (corrected p-value = standard genome-wide significance p-value/number of regions), and (ii) a “relaxed” threshold in keeping with prior work (Bernabeu et al., 2021) corresponding to the standard genome-wide significance threshold of p < 5e-8. For cortical phenotypes all measures in each category (regional CT, SA and GMV and corresponding global phenotypes) were grouped together for multiple testing correction resulting in p < 5e-8/(361) = 1.4e-10.

At the “strict” threshold, two SNPs showed statistically-significant sex differences in their phenotypic effects on regional cortical GMV measures (chr16:rs113078989 in BA6, chrX:rs747862348 in region Left TE1p; **Supplementary Table S14**), and none for CT and SA phenotypes (Figures 4A**, B, C**), or global and subcortical phenotypes. The SNPs underwent positional mapping (10 kb symmetric window) to protein coding genes in MAGMA (de Leeuw et al., 2015) as implemented in FUMA (Watanabe et al., 2017). The autosomal SNP chr16:rs113078989 mapped to the gene *RBFOX1* whereas chrX:rs747862348 did not map to any protein-coding genes. Figure 4D shows cortical regions where rs113078989 showed sex-biased relationships with GMV at the p < 0.05 level of significance, including the supplementary motor region (6MA), where this association also reached the strict statistical threshold (i.e. correction across the genome and all cortical regions). For 23 of these 26 cortical regions showing suggestive sex-biased association between rs113078989 and GMV (including region 6MA) - the sex-bias reflected a more prominent association in males than females (shown as a boxplot for 6MA in **Fig 4E**). This predominant male-bias in the effect of rs113078989 on GMV was also apparent when considering all cortical regions collectively (mean sex-difference z-scores of rs113078989 was positive on both hemispheres: mean z_left_= 0.55, mean z_right_= 0.15; Wilcoxon rank test p-values for mean z > 0: p_left_ = 7e-11, p_right_ = 0.04). Tissue specific expression (Fig. 4F) from GTEx data (V8, as implemented in FUMA) indicated high-expression in the brain for *RBFOX1* with additional muscular expression.

**Figure 4:**
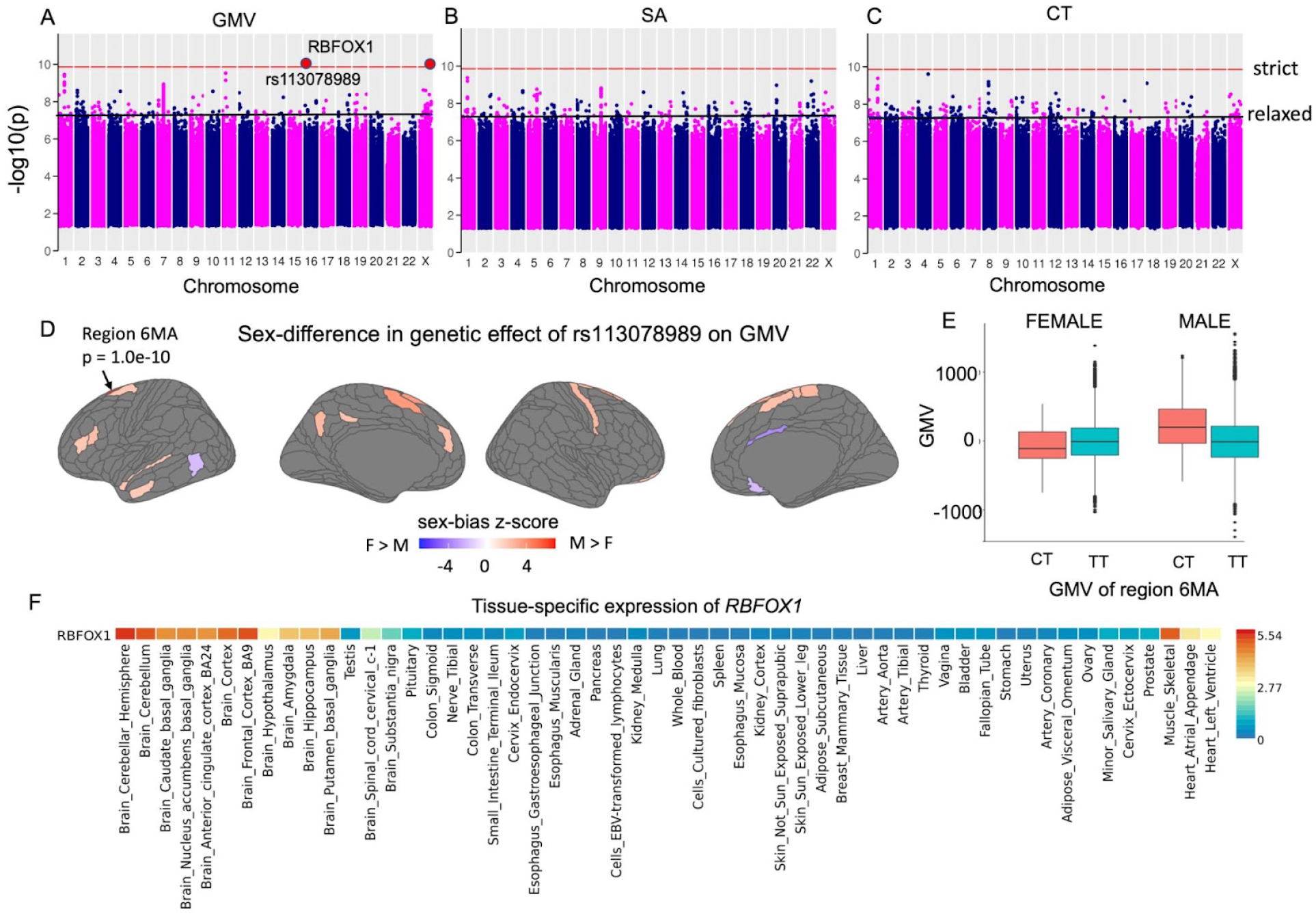
Genome-wide tests of sex-difference in SNP effects in regional GMV, SA and CT. (**A, B, C)**: minimum p-value across all 360 cortical regions for each SNP for each phenotype category: GMV (**A**), SA (**B**) and CT (**C**). The red and the blue horizontal lines correspond to the “strict” and the “relaxed” significance thresholds (p <1.38e-10 and p<5e-8, respectively). Red circles indicate SNPs above the “strict” threshold: chr16:rs113078989 and chrX:rs747862348. rs113078989 mapped to protein coding gene *RBFOX1*. **D**. Sex-difference in effect of rs113078989 across the cortex on cortical GMV shown as z-scores (**Methods**) with red indicating higher magnitude in males; all regions with sex-difference p-value > 0.05 are shown in gray. **E**: Boxplot showing GMV values for each sex for the CT and TT genotypes of rs113078989 in region 6MA . **F**: GTEx tissue expression heatmap plot for *RBFOX1* indicating significant brain expression (from FUMA).

SNPs passing the “relaxed” threshold mapped to 518 unique genes (99 SNPs mapped to 186 genes in GMV, 105 SNPs mapped to 157 genes in SA, 127 SNPS mapped to 178 genes in CT, 4 SNPS in subcortical volumes mapped to 13 genes, **Supplementary Tables S15-S19**). No SNP passed the “relaxed” threshold for the three global phenotypes. Functional analyses with GENE2FUNC in FUMA did not identify any statistically significant molecular function, biological process or cellular compartment GO term enrichments for genes identified using the relaxed threshold - regardless of whether mapped genes for each phenotype were considered as separate sets (i.e. regional cortical GMV, SA, CT and subcortical volumes), or combined into a unique gene set (comprising 336 independently significant SNPs which mapped to 505 genes) or whether the background gene set used for enrichment analysis was the default all gene option in FUMA or a set of brain-specific genes (Wagstyl et al., 2022).

## Discussion

Our study - which represents the first systematic survey for potential sex-differences in the common variant genetic architecture of human neuroanatomical variation - generated several findings of note, which are considered in turn below, along with important caveats and limitations.

First, we find that several previously reported observations regarding SNP-based h^2^ of brain anatomy when combining males and females (Elliott et al., 2018; Grasby et al., 2020; Hibar et al., 2015; Mallard et al., 2021; Satizabal et al., 2019; Smith et al., 2021) could be independently replicated in each sex group. Thus, within both males and females separately, we see that: mean cortical thickness is less heritable than total surface area or total GMV (Smith et al., 2021**; Supplementary Table S5**). We also find that in each sex some of the highest h^2^ values for regional cortical measures are seen in the primary and secondary visual cortex regions for GMV and SA. In CT we find highest h^2^ in the retrosplenial cortex in CT in males and in V2 in females. The high heritability of the visual cortex has been previously reported in both twin studies (Strike et al., 2018 reported highest genetic contribution to phenotypic variance in visual cortex SA) and population-based studies (Elliott et al., 2018; Grasby et al., 2020; Smith et al., 2021). The lowest values of h^2^ of GMV and SA are seen in areas including the rostral anterior cingulate cortex, medial prefrontal area, and frontal eye field - regions which are also reported to have some of the lowest h^2^ values in Smith et al., 2021. Similarly, of the 23 subcortical volumes examined, we find within both sexes that h^2^ values are highest for cerebellum, putamen and caudate nucleus and lowest for amygdala (**Supplementary Table S6**) consistent with findings of Satizabal et al. (2019) and Smith et al. (2021).

Second, although direct group comparisons did not identify any individual anatomical phenotype with statistically significant sex-differences in h^2^ estimates, we did find statistically significant differences in the distribution of regional h^2^ estimates for cortical GMV and SA, such that h^2^ estimates were on average higher for females than males. This tendency towards greater trait h^2^ in females has also been reported by some (Bernabeu et al., 2021) prior sex-stratified analysis of heritability of non-neuroanatomical traits (e.g. for systolic, diastolic blood pressure and waist circumference in (Ge et al., 2017; Gilks et al., 2014). Of the 31 traits considered in (Gilks et al., 2014), 15 showed higher h^2^ in females, 3 in males, and 15 showed no sex difference. We show that for the neuroanatomical phenotypes considered here, the observation of higher mean h^2^ in females is accompanied by higher mean regional V_G_ and V_P_ values in males than females. Thus, for the neuroanatomical phenotypes examined here, there is general tendency to greater phenotypic variance in males as compared to females, whereas the proportion of phenotypic variance accounted for by the additive effect of common variants is generally greater in females than males.

Third, since one of the motivations behind our work was the observed phenotypic sex-difference in brain phenotypes (e.g., Liu et al., 2020) we screened for significant association between phenotypic sex-difference and sex-difference in h^2^ as well as between-sex r_g_. We found no evidence of significant correlation between phenotypic sex-difference and sex-difference in h^2^ for GMV and SA, although a weak negative correlation was observed (r = -0.14, p = 0.007) for CT. Additionally, there was no statistical evidence for a general trend whereby regions with greater sex-differences in their mean values tend to show lower between sex genetic correlations in GMV or CT. However, a weak positive correlation was seen for SA (r = 0.20) with lower r_g_ corresponding to more female-biased regions. Also, both of the cortical phenotypes showing between-sex r_g_ values significantly below 1 also showed significant phenotypic sex differences in their means. Nevertheless, other than these few exceptions, there was little evidence for a strong or general trend for regions of greater sex-differences in phenotypic means showing larger sex differences in h^2^ or lower between-sex r_g_ values.

Fourth, in keeping with high between-sex r_g_ for almost all neuroanatomical phenotypes, we find very few SNPs (2 out of 12.7 million SNPS with MAF > 0.001) with sex-biased effects in GWAS after strict correction for multiple comparisons. With the aid of positional gene mapping (FUMA) we were able to map one of the SNPs (chr16:rs113078989) to *RBFOX1*-a synaptic gene expressed in both excitatory and inhibitory neurons (The human protein atlas, http://www.proteinatlas.org, Uhlén et al., 2015). *RBFOX1* encodes a splicing factor important for neuronal development and has been previously implicated in several male-biased neurodevelopmental and neuropsychiatric disorders including autism spectrum disorder (ASD), intellectual disability and epilepsy, attention-deficit hyperactivity disorder, schizoaffective disorder and schizophrenia (Cross-Disorder Group of the Psychiatric Genomics Consortium, 2019; Fogel et al., 2012). In our work, the *RBFOX1* implicating SNP rs113078989 shows male-specific effects on the GMV of region 6MA, part of the supplementary motor region known to play a role in coordinating complex movements. Male carriers of the minor allele (CT genotype) showed higher GMV compared to homozygous individuals (TT genotype). The fact that the sole gene implicated by these analyses is strongly associated with such sex-biased conditions is certainly striking and points towards ways in which sex-biased genetic effects could potentially shape sex differences in the prevalence or presentation of neurodevelopmental disorders. However, it will be crucial to test the replicability of this finding as larger datasets for sex-stratified GWAS of neuroimaging traits become available. Work in these datasets may also bring some of the subthreshold SNPs from our relaxed threshold analyses into statistical significance - potentially expanding the number of genetic variants with sex-biased influences on human brain development. Additionally, our finding of only two SNPs showing statistically significant sex-differentiated effects on brain anatomy is consistent with the recent suggestion by Zhu et al. (2022) that gene-by-sex interactions may largely act through sex differences in the magnitude of many genetic effects (“amplification”), rather than differences in the identity of causal variants or the direction of their effects.

Our findings should be considered in light of several caveats and limitations - some of which may be addressable in future research as datasets continue to increase in size and diversity. First, the UKB dataset - although revolutionary in its impact - predominantly includes individuals of European descent ages between 40 and 80 years. As such our findings cannot be assumed to generalize outside these demographic limits and it will be crucial to revisit the questions addressed in our current study within different phases of the lifespan and in populations with more diverse genetic ancestries. Second, our study design does not include rare single nucleotide variants with MAF <0.0001 (or MAF <0.001 for GWAS) or other classes of genetic variation such as indels or copy number variations - and future studies should also consider potentially sex-biased effects of these variant classes. Third, we have focused here on regional measures of brain anatomy using well-established parcellations of the human brain, but there are many alternative ways of measuring brain anatomy, and many other imaging derived phenotypes beyond those offered by structural MRI. Future studies should ideally extend to this broader range of phenotypes, although we note that the need for even more severe correction for multiple comparisons, and the lower measurement reproducibility for most imaging-derived phenotypes as compared to those structural MRI phenotypes we study here (Buimer et al. 2020; Knussmann et al. 2022; Noble et al. 2017), will substantially lower statistical power unless sample sizes are dramatically increased beyond those included here. Fourth, while there are strong theoretical grounds (and some preliminary empirical findings herein) to motivate continued comparison of genetic influences on brain anatomy between males and females - future studies should also consider sex differences in environmental influences on the brain, and consider the many partly dissociable aspects of sex and gender that we are to some extent obscuring by the necessary treatment of sex as a binary variable in the present study.

Notwithstanding the above caveats and limitations, our study provides the first tests for potential sex differences in the genetic architecture of human brain anatomy using one of the largest available individual level genotype and neuroimaging datasets. We investigate sex-differences in brain-linked genetic measures at the individual SNP-level as well as at the whole-genome level and find general concordance in the genetic basis of brain anatomical traits between males and females. Four notable exceptions to this general pattern are: 1) mean higher h^2^ in females for GMV and SA but not CT; 2) two cortical regional phenotypes showing detectable deviation from r_g_ = 1; 3) weak spatial correlations between sex-differences in anatomy and sex differences in h^2^ for CT, and lower r_g_ values for SA; and, 4) preliminary evidence for a sex-biased relationship between neuroanatomy and common genetic variation mapping to RBFOX1 - a gene implicated in the neurobiology of several sex-biased psychiatric disorders. The methods and results of this study - which represents the most systematic screen to date for sex differences in the genetic architecture of human neuroanatomical variation - offer a valuable reference point for the future studies to be undertaken as available datasets increase in sample size, diversity of genetic ancestry and phenotypic breadth.

## Data Availability

All data produced in the present study are available upon reasonable request to the authors

## Acknowledgement

This research was supported (in part) by the Intramural Research Program of the NIMH (ZIA MH002949-07, ZIC MH002960). TTM was supported by funds from NIH T32HG010464. This work utilized the computational resources of the NIH HPC Biowulf cluster. (http://hpc.nih.gov).

## Methods

### The UK biobank sample

Details of the UKB sample used in this work can be found in past studies (Mallard et al., 2021) and also at https://www.ukbiobank.ac.uk. We used the most recent release of brain MRI data (downloaded on April 23, 2020) for 38,685 samples together with the imputed genetic data provided by the UKB (Version 3) under application 22875. Our analyses included individuals with non-Hispanic European ancestry (according to UKB provided information) to avoid population stratification related confounding. After imaging and genetic quality control steps (as described below), the final data consisted of 14534 males and 16294 females (sample number varied with phenotype) and 17.38 million SNPs. All participants provided informed consent.

### Brain MRI phenotypes

Our study included regional cortical, regional subcortical and global brain phenotypes. T1w images were processed with FreeSurfer v.6.0.0 (Dale et al., 1999; Fischl et al., 2002; Fischl & Dale, 2000) to extract regional cortical gray matter volume (GMV), surface area (SA) and thickness (CT) using the multimodal HCP parcellation (Glasser et al., 2016), which divides each hemisphere into 180 regions. Mean cortical thickness, total cortical surface area and total brain volume were included as global measures. Lastly, 23 subcortical structure volumes (as calculated in the FreeSurfer pipeline) were included resulting in a total of 1106 phenotypes (**Supplementary Table S1**). Within each sex group these phenotypes were corrected for age, age^2^, head location in scanner, scanning site, head motion (calculated from resting fMRI, provided by UKB), and Euler number (Rosen et al., 2018) which is a measure of image quality. Regional cortical phenotypes were additionally corrected for respective global phenotypes: i.e., GMV for total brain volume, SA for total surface area, CT for mean cortical thickness and subcortical structure volumes for total brain volume.

### Quality control

MRI image quality control steps included removal of intracranial volume outliers (more than 4 SD away from the mean in each sex group) and samples with Euler number (reflecting image quality) < -217 (Rosen et al., 2018), resulting in a total of 30827 individuals (14534 males and 16294 females).

Genetic data consisted of imputed genotypes available from the UKB (Version 3). Data for individuals passing MRI quality control steps as described above were extracted using PLINK v2.0 (Purcell et al., 2007). Information provided by the UKB was used to remove individuals with putative sex chromosome aneuploidy, excessive heterozygosity, mismatched self-reported sex and genetic sex, and excessive relatedness. This was followed by removal of variants with imputation INFO score < 0.3, MAF < 0.0003, Hardy-Weinberg equilibrium p < 1e-6, missingness > 0.05, and variants with more than two alleles. For the X-chromosome non-pseudoautosomal (non-PAR) region, these filters were applied to males and females separately and variants passing quality control in both sexes were retained for analyses. Lastly, individuals with missingness > 0.1 and one person from each pair of individuals with relatedness > 0.05 (as described in next section) were also removed. The final set of genetic variants included 16.79 million autosomal and 585465 X-chromosome SNPs. For sex-stratified GWAS and sex-biased SNP effect calculation we used a more stringent MAF > 0.001 threshold (Elliott et al., 2018; Smith et al., 2021) resulting in a total of 12.1 million autosomal SNPs and 584816 X-chromosome SNPs. For the X-chromosome the MAF of SNPs was calculated separately for males and females and only SNP with MAF > 0.001 in both sexes were retained.

For each category (GMV, SA and CT) of regional cortical phenotype, only individuals with all phenotypes (all 360 regions) within 5 SD of the mean were included in the genetic analyses resulting in 14140 males and 15846 females for GMV, 14271 males and 15899 females for SA and 14301 males and 16054 females for CT.

### SNP-heritability, genetic correlation, and genome-wide association

Sex-specific SNP-based heritability (h^2^) of each phenotype was estimated using sex-specific genomic relatedness matrices (GRMs) in GCTA (Yang et al., 2011) reflecting genetic similarity between pairs of individuals. GRMs were constructed for the autosomes and the X-chromosome separately in GCTA v 1.93 (Yang et al., 2011). To ensure closely related individuals were not included in the analyses, the autosomal GRM was used to exclude one individual from each pair with relatedness > 0.05. The total SNP-heritability for each phenotype in each sex was estimated using a joint model including both the autosomal and X-chromosome GRMs. P-values corresponding to the significance of h^2^ were estimated using likelihood ratio tests implemented in GCTA. The first 10 genetic ancestry principal components were included as covariates for the heritability analyses. For each phenotype X-linked h^2^ was estimated using the best-fit dosage compensation model from (Mallard et al., 2021).

For a trait, genetic correlation r_g_ between the sexes was estimated also using GCTA. Since r_g_ estimation can be unstable for low trait heritability, only traits with h^2^ p-value < 0.05 were used. For this a modified GRM approach was used following (Yang et al., 2015): autosomal GRMs were calculated combining the males and females and modified phenotype files with two columns corresponding to the male and the female phenotypes were used with the “--reml-bivar” option. Deviation of r_g_ from 1 was estimated using the “--reml-bivar-rg 1” option. We did not estimate r_g_ for the X-chromosome due to lack of statistical power (the highest h^2^_X-chr_ in GMV was 0.034+/-0.02 in males). For r_g_ calculation also the first 10 ancestry PCs were used as covariates. Sex-stratified genome-wide association analyses (GWA - encompassing autosomes and the X-chromosome non-PAR region) for each phenotype were performed in PLINK V2.0 (Purcell et al., 2007) using the --linear option and the 10 ancestry PCs as covariates. Cortical surface plots were created using the “ggseg” package (Mowinckel & Vidal-Piñeiro, 2020) in R (R Core Team, 2021).

### Estimating sex-differences

For each phenotype sex difference in total h^2^ was estimated by calculating the following z-score:

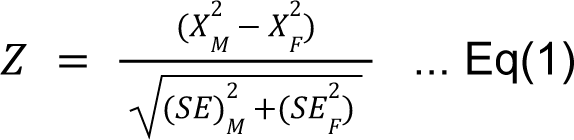

where X^2^ and X^2^ are male and female h^2^ and SE and SE are the standard errors of the respective h^2^ estimates (similar to the approach of (Martin et al., 2021). Corresponding p-values were then calculated as p = 2 × (1 - Φ(|*Z*|)), where Φ is the cumulative distribution function of the standard normal distribution. For the regional cortical phenotypes we report any result as significant for p < 1.4e-4, correcting for the number of cortical regions. For subcortical structures, we use a significance threshold of p < 0.0021 to correct for the 23 volumes tested.

Sex-diffrences in V_G_, V_P_, and V_E_ (additive genetic, phenotypic and environmental variance, respectively) were tested using paired t.tests as well as Wilcoxon rank test in R. Phenotypic sex-difference was estimated by testing the significance of the coefficient “b” of “sex” in the linear model (“lm” function in R):

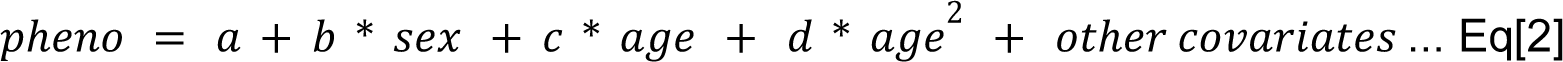

where “pheno” corresponds to a GMV, SA or CT phenotype and “other covariates” are described earlier in Methods. The potential sex-difference in the relationship between V_P_ and V_G_ was explored (**Figure S4, Supplementary Table S8**) by fitting the data to two models (moving to the simpler Eq [4] in the absence of evidence for significant quadratic effects from Eq [3] shown below and testing significance of the coefficients “d” and “f”:

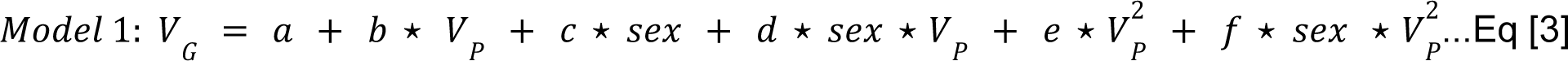

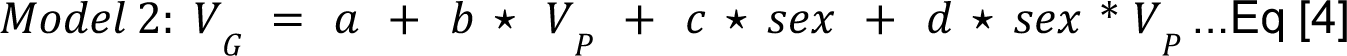

Sex-bias in SNP effect size was estimated in a similar way for each variant and each phenotype using Eq [1] where X^2^ and X^2^ represented male and female GWAS effect sizes for that phenotype and SE_M_ and SE_F_ represented the corresponding standard errors of the effect sizes. The p-values calculated from these z-scores represented the significance of sex-bias in SNP effects. We set two thresholds for downstream analyses: (i) a “strict” threshold correcting for multiple phenotypes by setting significance threshold to p < 1.4e-10, and (ii) a “relaxed” threshold using the standard genome-wide significance level of p < 5e-8. For subcortical structures we set the “strict” threshold to be p < 2.17e-9 corresponding to correction for 23 phenotypes.

### Gene mapping and functional analyses

SNPs with significant (strict or relaxed) sex-bias in effect sizes were mapped to genes using SNP2GENE in FUMA (Watanabe et al., 2017). Minor allele frequencies and LD structures were calculated in FUMA using the 1000 Genome phase 3 EUR population (1000 Genomes Project Consortium et al., 2015). First, the input SNPs were filtered for independent SNPs with r^2^ < 0.6. For each independent SNP, all known SNPs with r^2^>0.6 with one of the independent significant SNPs were included for further analyses (candidate SNPs). Based on the identified independent significant SNPs, independent lead SNPs were defined if they were independent from each other at r^2^ < 0.1. Additionally, if LD blocks of independent significant SNPs were closely located to each other (<250 kb), they were merged into one genomic locus. Each genomic locus could thus contain multiple independent significant SNPs and lead SNPs.

Using a positional mapping strategy (10 kb symmetric window) candidate SNPs were mapped to genes in SNP2GENE using MAGMA (de Leeuw et al., 2015). These genes were next used in GENE2FUNC with a background of 16573 brain expressed genes (Wagstyl et al., 2022) to test for gene set enrichment of the various GO categories (10532 gene sets in MsigDB V7.0 for molecular function, cellular components, biological function). Bonferroni-corrected p-value threshold for this step was set to 0.05/10532 = 4.7e-6.

## References

1. 1000 Genomes Project Consortium, Auton A., Brooks L.D., Durbin R.M., et al. (2015). A global reference for human genetic variation. Nature, 526(7571).

2. Bernabeu, E., Canela-Xandri, O., Rawlik, K., Talenti, A., Prendergast, J., & Tenesa, A. (2021). Sex differences in genetic architecture in the UK Biobank. Nature Genetics, 53(9), 1283–1289.

3. Buimer, E. E. L., Pas, P., Brouwer, R. M., Froeling, M., Hoogduin, H., Leemans, A., Luijten, P., van Nierop, B. J., Raemaekers, M., Schnack, H. G., Teeuw, J., Vink, M., Visser, F., Hulshoff Pol, H. E., & Mandl, R. C. W. (2020). The YOUth cohort study: MRI protocol and test-retest reliability in adults. Developmental Cognitive Neuroscience, 45, 100816.

4. Corre, C., Friedel, M., Vousden, D. A., Metcalf, A., Spring, S., Qiu, L. R., Lerch, J. P., & Palmert, M. R. (2016). Separate effects of sex hormones and sex chromosomes on brain structure and function revealed by high-resolution magnetic resonance imaging and spatial navigation assessment of the Four Core Genotype mouse model. Brain Structure & Function, 221(2), 997–1016.

5. Cross-Disorder Group of the Psychiatric Genomics Consortium (2019). Genomic Relationships, Novel Loci, and Pleiotropic Mechanisms across Eight Psychiatric Disorders. Cell, 179(7), 1469–1482.e11.

6. Dale, A. M., Fischl, B., & Sereno, M. I. (1999). Cortical surface-based analysis. I. Segmentation and surface reconstruction. NeuroImage, 9(2), 179–194.

7. DeCasien, A. R., Guma, E., Liu, S., & Raznahan, A. (2022). Sex differences in the human brain: a roadmap for more careful analysis and interpretation of a biological reality. Biology of Sex Differences, 13(1), 43.

8. de Leeuw, C. A., Mooij, J. M., Heskes, T., & Posthuma, D. (2015). MAGMA: generalized gene-set analysis of GWAS data. PLoS Computational Biology, 11(4), e1004219.

9. Elliott, L. T., Sharp, K., Alfaro-Almagro, F., Shi, S., Miller, K. L., Douaud, G., Marchini, J., & Smith, S. M. (2018). Genome-wide association studies of brain imaging phenotypes in UK Biobank. Nature, 562(7726), 210–216.

10. Fischl, B., & Dale, A. M. (2000). Measuring the thickness of the human cerebral cortex from magnetic resonance images. Proceedings of the National Academy of Sciences, 97(20), 11050–11055.

11. Fischl, B., Salat, D. H., Busa, E., Albert, M., Dieterich, M., Haselgrove, C., van der Kouwe, A., Killiany, R., Kennedy, D., Klaveness, S., Montillo, A., Makris, N., Rosen, B., & Dale, A. M. (2002). Whole brain segmentation: automated labeling of neuroanatomical structures in the human brain. Neuron, 33(3), 341–355.

12. Fogel, B. L., Wexler, E., Wahnich, A., Friedrich, T., Vijayendran, C., Gao, F., Parikshak, N., Konopka, G., & Geschwind, D. H. (2012). RBFOX1 regulates both splicing and transcriptional networks in human neuronal development. Human Molecular Genetics, 21(19), 4171–4186.

13. Ge, T., Chen, C.-Y., Neale, B. M., Sabuncu, M. R., & Smoller, J. W. (2017). Phenome-wide heritability analysis of the UK Biobank. PLoS Genetics, 13(4), e1006711.

14. Gilks, W. P., Abbott, J. K., & Morrow, E. H. (2014). Sex differences in disease genetics: evidence, evolution, and detection. Trends in Genetics: TIG, 30(10), 453–463.

15. Glasser, M. F., Coalson, T. S., Robinson, E. C., Hacker, C. D., Harwell, J., Yacoub, E., Ugurbil, K., Andersson, J., Beckmann, C. F., Jenkinson, M., Smith, S. M., & Van Essen, D. C. (2016). A multi-modal parcellation of human cerebral cortex. Nature, 536, 171.

16. Grasby, K. L., Jahanshad, N., Painter, J. N., Colodro-Conde, L., Bralten, J., Hibar, D. P., Lind, P. A., Pizzagalli, F., Ching, C. R. K., McMahon, M. A. B., Shatokhina, N., Zsembik, L. C. P., Thomopoulos, S. I., Zhu, A. H., Strike, L. T., Agartz, I., Alhusaini, S., Almeida, M. A. A., Alnæs, D., … Null, N. (2020). The genetic architecture of the human cerebral cortex. Science, 367(6484).

17. Guma, E., Beauchamp, A., Liu, S., Levitis, E., Clasen, L. S., Torres, E., Blumenthal, J., Lalonde, F., Qiu, L. R., Hrncir, H., MacKenzie-Graham, A., Yang, X., Arnold, A. P., Lerch, J. P., & Raznahan, A. (2022). A cross-species study of sex chromosome dosage effects on mammalian brain anatomy. bioRxiv. https://doi.org/10.1101/2022.08.30.505916

18. Guma, E., Devenyi, G. A., Malla, A., Shah, J., Chakravarty, M. M., & Pruessner, M. (2017). Neuroanatomical and Symptomatic Sex Differences in Individuals at Clinical High Risk for Psychosis. Frontiers in Psychiatry / Frontiers Research Foundation, 8, 291.

19. Hibar, D. P., Stein, J. L., Renteria, M. E., Arias-Vasquez, A., Desrivières, S., Jahanshad, N., Toro, R., Wittfeld, K., Abramovic, L., Andersson, M., Aribisala, B. S., Armstrong, N. J., Bernard, M., Bohlken, M. M., Boks, M. P., Bralten, J., Brown, A. A., Chakravarty, M. M., Chen, Q., … Medland, S. E. (2015). Common genetic variants influence human subcortical brain structures. Nature, 520(7546), 224–229.

20. Knussmann, G. N., Anderson, J. S., Prigge, M. B. D., Dean, D. C., 3rd, Lange, N., Bigler, E. D., Alexander, A. L., Lainhart, J. E., Zielinski, B. A., & King, J. B. (2022). Test-retest reliability of FreeSurfer-derived volume, area and cortical thickness from MPRAGE and MP2RAGE brain MRI images. Neuroimage. Reports, 2(2). https://doi.org/10.1016/j.ynirp.2022.100086

21. Liu, S., Seidlitz, J., Blumenthal, J. D., Clasen, L. S., & Raznahan, A. (2020). Integrative structural, functional, and transcriptomic analyses of sex-biased brain organization in humans. Proceedings of the National Academy of Sciences of the United States of America, 117(31), 18788–18798.

22. Lotze, M., Domin, M., Gerlach, F. H., Gaser, C., Lueders, E., Schmidt, C. O., & Neumann, N. (2019). Novel findings from 2,838 Adult Brains on Sex Differences in Gray Matter Brain Volume. Scientific Reports, 9(1), 1671.

23. Mallard, T. T., Liu, S., Seidlitz, J., Ma, Z., Moraczewski, D., Thomas, A., & Raznahan, A. (2021). X-chromosome influences on neuroanatomical variation in humans. Nature Neuroscience, 24(9), 1216–1224.

24. Martin, J., Khramtsova, E. A., Goleva, S. B., Blokland, G. A. M., Traglia, M., Walters, R. K., Hübel, C., Coleman, J. R. I., Breen, G., Børglum, A. D., Demontis, D., Grove, J., Werge, T., Bralten, J., Bulik, C. M., Lee, P. H., Mathews, C. A., Peterson, R. E., Winham, S. J., … Sex Differences Cross-Disorder Analysis Group of the Psychiatric Genomics Consortium. (2021). Examining Sex-Differentiated Genetic Effects Across Neuropsychiatric and Behavioral Traits. Biological Psychiatry, 89(12), 1127–1137.

25. McCarthy, M. M. (2020). A new view of sexual differentiation of mammalian brain. Journal of Comparative Physiology. A, Neuroethology, Sensory, Neural, and Behavioral Physiology, 206(3), 369–378.

26. McCarthy, M. M., Arnold, A. P., Ball, G. F., Blaustein, J. D., & De Vries, G. J. (2012). Sex differences in the brain: the not so inconvenient truth. The Journal of Neuroscience: The Official Journal of the Society for Neuroscience, 32(7), 2241–2247.

27. McCarthy, M. M., Nugent, B. M., & Lenz, K. M. (2017). Neuroimmunology and neuroepigenetics in the establishment of sex differences in the brain. Nature Reviews. Neuroscience, 18(8), 471–484.

28. Mowinckel, A. M., & Vidal-Piñeiro, D. (2020). Visualization of Brain Statistics With R Packages ggseg and ggseg3d. Advances in Methods and Practices in Psychological Science, 3(4), 466–483.

29. Neufang, S., Specht, K., Hausmann, M., Güntürkün, O., Herpertz-Dahlmann, B., Fink, G. R., & Konrad, K. (2008). Sex Differences and the Impact of Steroid Hormones on the Developing Human Brain. Cerebral Cortex, 19(2), 464–473.

30. Noble, S., Spann, M. N., Tokoglu, F., Shen, X., Constable, R. T., & Scheinost, D. (2017). Influences on the Test–Retest Reliability of Functional Connectivity MRI and its Relationship with Behavioral Utility. Cerebral Cortex, 27(11), 5415–5429.

31. Premachandran, H., Zhao, M., & Arruda-Carvalho, M. (2020). Sex Differences in the Development of the Rodent Corticolimbic System. Frontiers in Neuroscience, 14, 583477.

32. Purcell, S., Neale, B., Todd-Brown, K., Thomas, L., Ferreira, M. A. R., Bender, D., Maller, J., Sklar, P., de Bakker, P. I. W., Daly, M. J., & Sham, P. C. (2007). PLINK: a tool set for whole-genome association and population-based linkage analyses. American Journal of Human Genetics, 81(3), 559–575.

33. R Core Team (2021). R: A language and environment for statistical computing. R Foundation for Statistical Computing, Vienna, Austria. URL https://www.R-project.org/.

34. Ritchie, S. J., Cox, S. R., Shen, X., Lombardo, M. V., Reus, L. M., Alloza, C., Harris, M. A., Alderson, H. L., Hunter, S., Neilson, E., Liewald, D. C. M., Auyeung, B., Whalley, H. C., Lawrie, S. M., Gale, C. R., Bastin, M. E., McIntosh, A. M., & Deary, I. J. (2018). Sex Differences in the Adult Human Brain: Evidence from 5216 UK Biobank Participants. Cerebral Cortex, 28(8), 2959–2975.

35. Rosen, A. F. G., Roalf, D. R., Ruparel, K., Blake, J., Seelaus, K., Villa, L. P., Ciric, R., Cook, P. A., Davatzikos, C., Elliott, M. A., Garcia de La Garza, A., Gennatas, E. D., Quarmley, M., Schmitt, J. E., Shinohara, R. T., Tisdall, M. D., Craddock, R. C., Gur, R. E., Gur, R. C., & Satterthwaite, T. D. (2018). Quantitative assessment of structural image quality. NeuroImage, 169, 407–418.

36. Satizabal, C. L., Adams, H. H. H., Hibar, D. P., White, C. C., Knol, M. J., Stein, J. L., Scholz, M., Sargurupremraj, M., Jahanshad, N., Roshchupkin, G. V., Smith, A. V., Bis, J. C., Jian, X., Luciano, M., Hofer, E., Teumer, A., van der Lee, S. J., Yang, J., Yanek, L. R., … Ikram, M. A. (2019). Genetic architecture of subcortical brain structures in 38,851 individuals. Nature Genetics, 51(11), 1624–1636.

37. Smith, S. M., Douaud, G., Chen, W., Hanayik, T., Alfaro-Almagro, F., Sharp, K., & Elliott, L. T. (2021). An expanded set of genome-wide association studies of brain imaging phenotypes in UK Biobank. Nature Neuroscience, 24(5), 737–745.

38. Strike, L. T., Hansell, N. K., Couvy-Duchesne, B., Thompson, P. M., de Zubicaray, G. I., McMahon, K. L., & Wright, M. J. (2018). Genetic Complexity of Cortical Structure: Differences in Genetic and Environmental Factors Influencing Cortical Surface Area and Thickness. Cerebral Cortex, 29(3), 952–962.

39. Sudlow, C., Gallacher, J., Allen, N., Beral, V., Burton, P., Danesh, J., Downey, P., Elliott, P., Green, J., Landray, M., Liu, B., Matthews, P., Ong, G., Pell, J., Silman, A., Young, A., Sprosen, T., Peakman, T., & Collins, R. (2015). UK biobank: an open access resource for identifying the causes of a wide range of complex diseases of middle and old age. PLoS Medicine, 12(3), e1001779.

40. Supekar, K., de Los Angeles, C., Ryali, S., Cao, K., Ma, T., & Menon, V. (2022). Deep learning identifies robust gender differences in functional brain organization and their dissociable links to clinical symptoms in autism. The British Journal of Psychiatry: The Journal of Mental Science, 1–8.

41. Thompson, P. (2020). ENIGMA and Global Neuroscience: A Decade of Large-Scale Studies of the Brain in Health and Disease Across More Than 40 Countries. In Biological Psychiatry (Vol. 87, Issue 9, p. S56). https://doi.org/10.1016/j.biopsych.2020.02.167

42. Uhlén, M., Fagerberg, L., Hallström, B. M., Lindskog, C., Oksvold, P., Mardinoglu, A., Sivertsson, Å., Kampf, C., Sjöstedt, E., Asplund, A., Olsson, I., Edlund, K., Lundberg, E., Navani, S., Szigyarto, C. A.-K., Odeberg, J., Djureinovic, D., Takanen, J. O., Hober, S., … Pontén, F. (2015). Tissue-based map of the human proteome. Science, 347(6220), 1260419.

43. Visscher, P. M., Wray, N. R., Zhang, Q., Sklar, P., McCarthy, M. I., Brown, M. A., & Yang, J. (2017). 10 Years of GWAS Discovery: Biology, Function, and Translation. American Journal of Human Genetics, 101(1), 5–22.

44. Vousden, D. A., Corre, C., Spring, S., Qiu, L. R., Metcalf, A., Cox, E., Lerch, J. P., & Palmert, M. R. (2018). Impact of X/Y genes and sex hormones on mouse neuroanatomy. NeuroImage, 173, 551–563.

45. Wagstyl, K., Adler, S., Seidlitz, J., Vandekar, S., Mallard, T. T., Dear, R., DeCasien, A. R., Satterthwaite, T. D., Liu, S., Vértes, P. E., Shinohara, R. T., Alexander-Bloch, A., Geschwind, D. H., & Raznahan, A. (2022). Transcriptional Cartography Integrates Multiscale Biology of the Human Cortex. In bioRxiv (p. 2022.06.13.495984). https://doi.org/10.1101/2022.06.13.495984.

46. Warling, A., Yavi, M., Clasen, L. S., Blumenthal, J. D., Lalonde, F. M., Raznahan, A., & Liu, S. (2021). Sex Chromosome Dosage Effects on White Matter Structure in the Human Brain. Cerebral Cortex, 31(12), 5339–5353.

47. Watanabe, K., Taskesen, E., van Bochoven, A., & Posthuma, D. (2017). Functional mapping and annotation of genetic associations with FUMA. Nature Communications, 8(1), 1826.

48. Williams, C. M., Peyre, H., Toro, R., & Ramus, F. (2021a). Neuroanatomical norms in the UK Biobank: The impact of allometric scaling, sex, and age. Human Brain Mapping, 42(14), 4623–4642.

49. Williams, C. M., Peyre, H., Toro, R., & Ramus, F. (2021b). Sex differences in the brain are not reduced to differences in body size. Neuroscience and Biobehavioral Reviews, 130, 509–511.

50. Wright, C. L., Schwarz, J. S., Dean, S. L., & McCarthy, M. M. (2010). Cellular mechanisms of estradiol-mediated sexual differentiation of the brain. Trends in Endocrinology and Metabolism: TEM, 21(9), 553–561.

51. Yang, J., Bakshi, A., Zhu, Z., Hemani, G., Vinkhuyzen, A. A. E., Nolte, I. M., van Vliet-Ostaptchouk, J. V., Snieder, H., Lifelines Cohort Study, Esko, T., Milani, L., Mägi, R., Metspalu, A., Hamsten, A., Magnusson, P. K. E., Pedersen, N. L., Ingelsson, E., & Visscher, P. M. (2015). Genome-wide genetic homogeneity between sexes and populations for human height and body mass index. Human Molecular Genetics, 24(25), 7445–7449.

52. Yang, J., Lee, S. H., Goddard, M. E., & Visscher, P. M. (2011). GCTA: a tool for genome-wide complex trait analysis. American Journal of Human Genetics, 88(1), 76–82.

53. Zhao, B., Ibrahim, J. G., Li, Y., Li, T., Wang, Y., Shan, Y., Zhu, Z., Zhou, F., Zhang, J., Huang, C., Liao, H., Yang, L., Thompson, P. M., & Zhu, H. (4865). “Heritability of regional brain volumes in large-scale neuroimaging and genetic studies”: Corrigendum. Cerebral Cortex, 31(10). https://doi.org/10.1093/cercor/bhab270

