## Supplementary Figures for "A sex-stratified analysis of the genetic architecture of human brain anatomy"

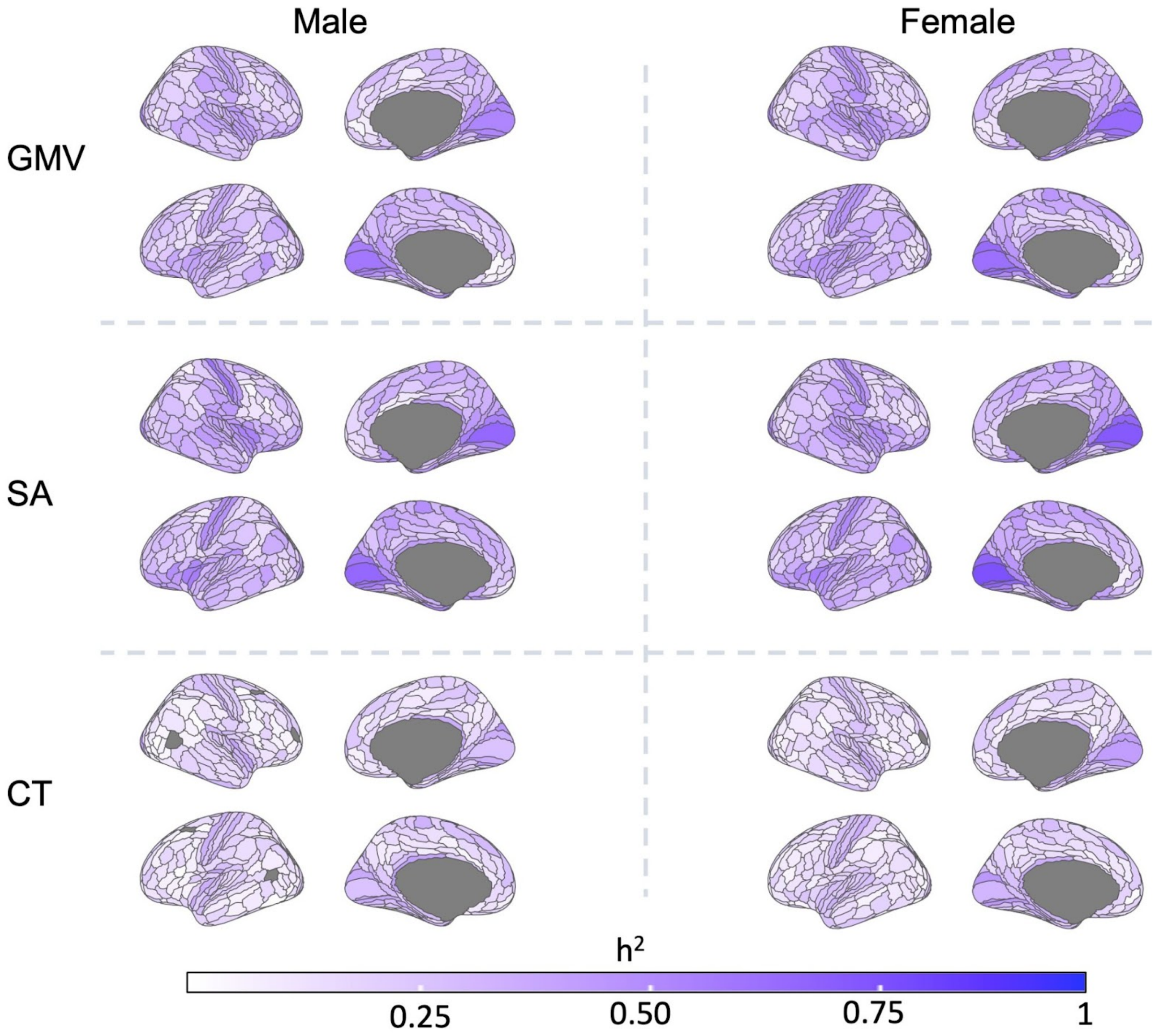


**Figure S1**: **SNP-based heritability ($h^2$) of GMV, SA and CT in the UKB**. Similar to **Figure 2** in the main text (but for both hemispheres).

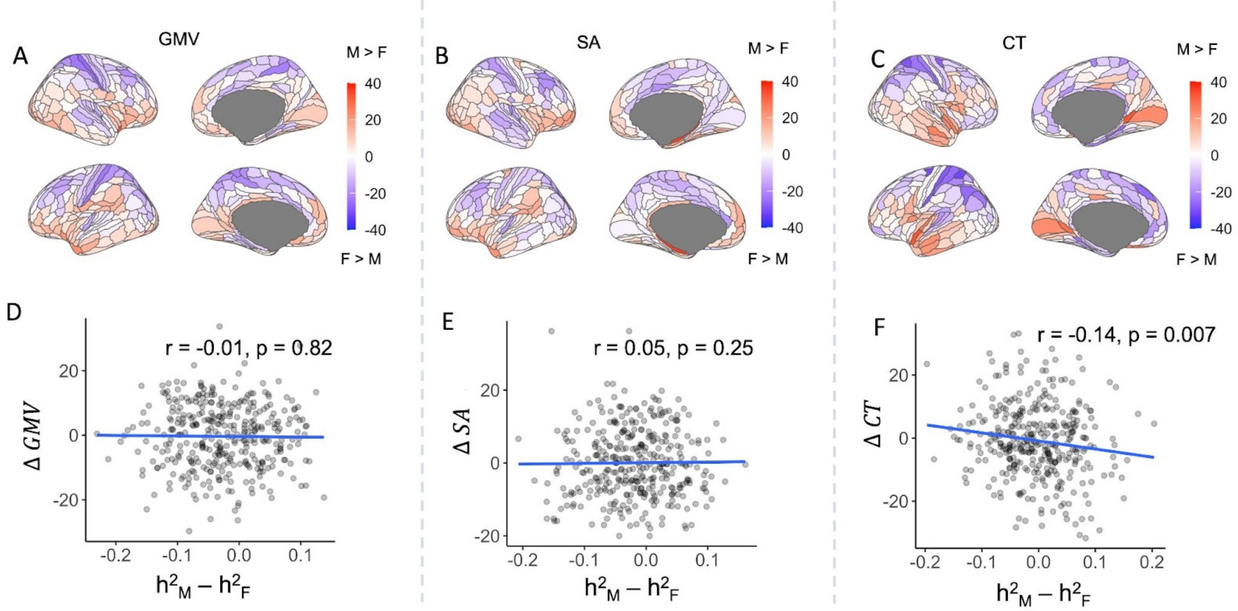


**Figure S2**: **Comparison between phenotypic sex difference and sex difference in $h^2$**. (**A**, **B**, **C)**: Phenotypic sex differences for GMV (**A**), SA (**B**) and CT (**C**) shown as t-statistics of the coefficient of the "sex" term in a linear model which also controlled for age, $age^2$, scanner position, Euler number, scan center and corresponding global phenotypes (**Methods**). Red indicates male bias whereas blue indicates female bias. (**D**, **E**, **F)**: scatter plots of sex-difference in $h^2$ (male - female) vs. phenotypic sex-difference for GMV (**D**), SA (**E**) and CT (**F**). In these plots a positive coordinate indicates male bias.

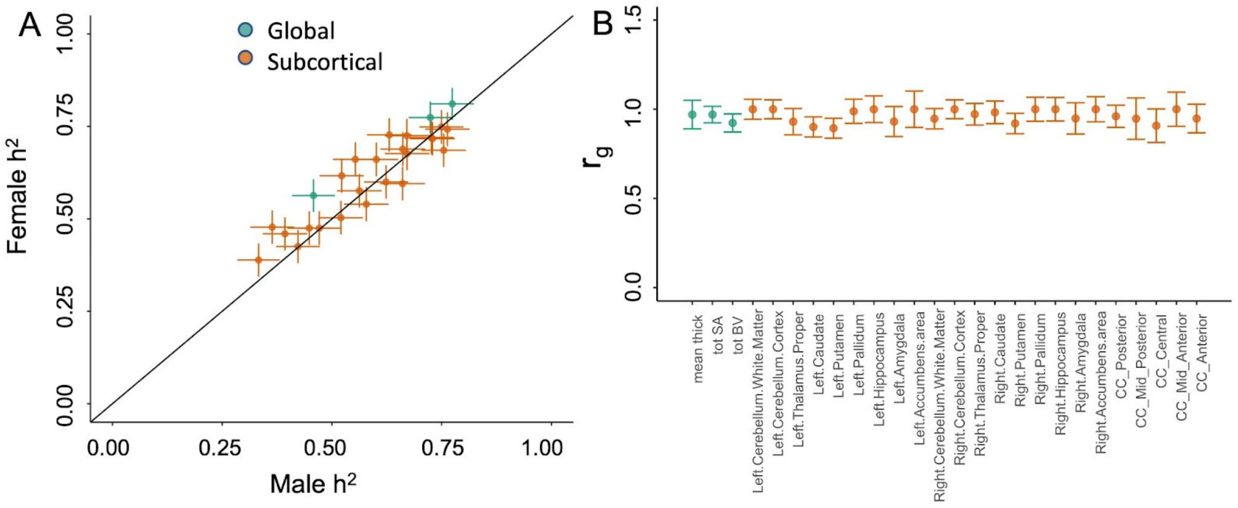


**Figure S3: Heritability and genetic correlation in subcortical and global phenotypes**. **A**: Scatter plot showing male and female $h^2$ for 3 global and 23 subcortical volumes. **B**: Autosomal genetic correlation ($r_g$) between males and females for the same phenotypes as calculated by GCTA.

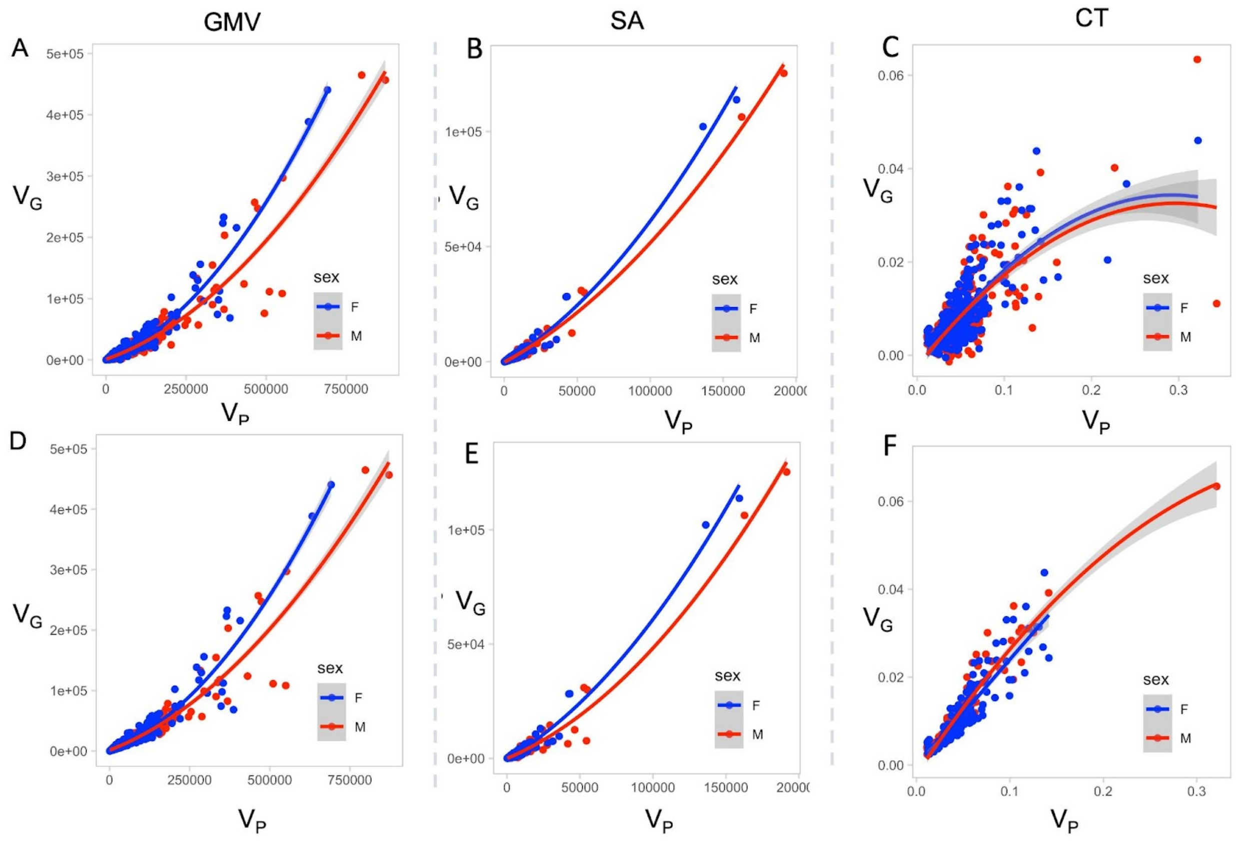


**Figure S4**: **Scatter plots comparing genetic variance $V_G$ and phenotypic variance $V_p$ in both sexes**. **A**, **B**, **C** show these values for each region in the HCP parcellation (180 regions in each hemisphere) for GMV, SA and CT corrected for age, age$^2$, scanner position, Euler number, scan center and corresponding global phenotypes. **D**, **E**, **F** show similar plots but for regions which showed significant $h^2$ ($p < 1.4e\text{-}4$) only. The blue and red solid lines show sex-specific fits to the data for which the coefficients can be found in Table S8.
